# Psychological and social risk factors and pregnancy outcome: a prospective cohort study

**DOI:** 10.1101/2024.01.14.24301280

**Authors:** Elbert-Jaap I. Schipper, Klaas J. Wardenaar, Annemieke C. Bolte, Rei Monden, Titus W.D.P. van Os, Marieke Wichers, Peter de Jonge

## Abstract

**Background:** Research suggests that besides somatic factors, psychological and social factors are associated with pregnancy outcomes. The objective of this study was to determine the independent effects of psychosocial risk factors on maternal blood pressure, gestational age at birth, birthweight and Apgar score.

**Methods:** In a prospective cohort study, Dutch women in the first half of pregnancy were recruited at a regional hospital, a university hospital and ten midwife practices. At inclusion, participants (before 20 weeks of gestation) filled in questionnaires on social and psychological factors. Obstetric data were extracted from patient records. Associations between risk factors and pregnancy outcomes were analysed with multivariable linear and logistic regression.

**Results:** 598 Women were included in the study. 14 Women did not return questionnaires and 12 women stopped study participation before delivery. In multivariable regression analysis, primiparity, Odds Ratio [OR] = 3.4 (1.5, 7.5), obstetric and somatic history, OR = 3.5 (1.5, 7.9), and diastolic blood pressure at intake, OR = 1.1 (1.0, 1.1), were independently associated with preterm delivery. Smoking status, OR = 5.5 (2.3, 13), was independently associated with a newborn small for gestational age, and primiparity, OR = 6.9 (1.1, 45), with a low Apgar score. Diastolic blood pressure at intake, OR = 1.1 (1.07, 1.14), hypertension at intake, OR = 3.6 (1.1, 11), and negative affect, OR = 1.1 (1.02, 1.14), were independently associated with gestational hypertension. Negative affect was the only psychosocial risk factor independently associated with pregnancy outcome.

**Conclusion:** Although psychosocial factors are important in obstetric care, measurement of these factors in early pregnancy seems to have limited independent predictive value for adverse pregnancy outcome when medical and/or obstetric history and commonly applied physical measurements are already considered.

## Background

Pregnancy complications have profound emotional impact and form a major public health concern.^1^ Successful pregnancy demands specific conditions in the maternal body, where the vascular and immunological systems are pivotal. Diseases involving these systems (i.e. hypertension and autoimmune disease) are well-known risk factors for complications of pregnancy.^2,3^

Besides somatic health, psychological and social factors have also been observed to be associated with pregnancy outcome. This could be explained by hormonal and immunological disturbances in depression, anxiety and stress, and by unhealthy behaviours (e.g. substance abuse).^4^ Maternal depression has been linked to increased risk of preeclampsia, preterm delivery and low birthweight.^5,6^ Preterm delivery and low birthweight have also been found to be associated with maternal anxiety symptoms,^7^ stressful live events and living in a deprived neighborhood.^4,8^ Depression and anxiety are the most frequently studied psychological phenomena in pregnancy. However, other characteristics like personality and attachment style might also be important as they have been shown to be correlated with other somatic conditions (e.g. hypertension, stroke).^9,10^ Indeed, available publications indicate that high neuroticism, low extraversion and insecure attachment increase the probability of pregnancy complications.^11,12^ Psychosocial risk factors for pregnancy complications correlate with each other and with somatic risk factors, which obscures their independent roles in pathological pathways.^13^

In obstetric practice several known risk factors are routinely assessed, including medical and obstetric history, body mass index (BMI), blood pressure and smoking status. Screening for anxiety and depression is increasingly advised, mainly because of adverse effects of these conditions on mental development of the foetus and the newborn.^13,14^ Timely treatment of the mother’s depressive and/or anxiety disorder may prevent these negative effects.^15^ Knowing that psychosocial factors are also correlated with pregnancy complications raises the question if adding these factors to screening procedures could improve early prediction of complications. This could easily be implemented, using questionnaires. In the current study we tested such a broad screening procedure in an integrative prospective design, to assess the added value of measuring social and psychological risk factors before 20 weeks gestation.

## Methods

### Design

In a prospective cohort study, Dutch women in the first half of pregnancy were recruited at a regional hospital (MCL Leeuwarden), a university hospital (VU University Medical Centre Amsterdam) and ten midwife practices in these regions from January 2007 until November 2010. Inclusion criteria were singleton pregnancy, age over 17 years and sufficient command of the Dutch language. At baseline (before 20 weeks of gestation), participants filled in questionnaires assessing body height, smoking status and social and psychological factors (see below). From the obstetric records we extracted data on somatic disease, course of previous pregnancies and current pregnancy, maternal age, bodyweight, blood pressure and medication use. Figure 1 depicts a flow chart of inclusion. 467 Women (43.8%) declined participation. Reasons most frequently given for not participating were not having time or not feeling like it. Of 572 participants, questionnaires and delivery data were obtained.

**Figure 1.**
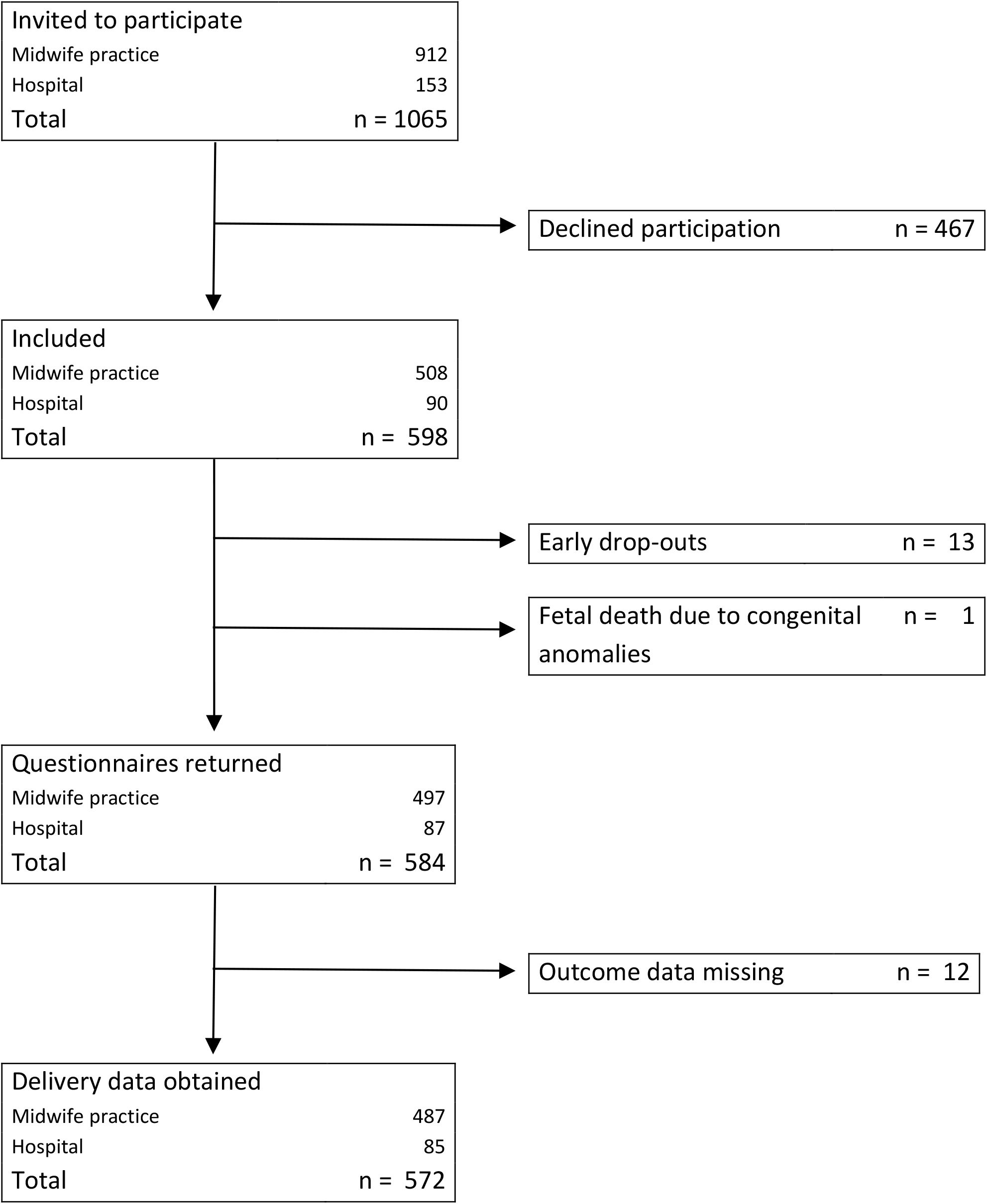
Flow chart of inclusion

### Questionnaires

To assess personality, the 12-item neuroticism and extraversion scales of the Eysenck Personality Questionnaire Revised Short Scale (EPQ-RSS)^16^ were administered. These scales both have high Cronbach’s α coefficients of 0.86 and 0.85, respectively.^16^

The Relationship Questionnaire (RQ) measures attachment style, a person’s pattern of interactions in relationships.^17^ The questionnaire consists of descriptions of each of four attachment styles (secure, dismissing, preoccupied and fearful) and respondents rate the extent to which each applies to them using a 1-7 Likert response scale. Two RQ scores are computed for image of self and of others.^18^

The 14-item Type D scale-14 (DS14) measures the personality traits of negative affectivity (the tendency to experience negative emotions) and social inhibition (the tendency to suppress the expression of emotions in social interaction).^19^ The instrument comprises statements (7 per trait) about oneself, which respondents rate on a 0 (“false”) - 4 (“true”) Likert scale. Two sum scores are computed (negative affectivity and social inhibition), with high internal consistency in a previous study (α=0.88 and α=0.86, respectively).^19^

A 23-item version of the List of Threatening Experiences (LTE) was administered to assess the occurrence of adverse life events in the previous six months.^20^ The 23-item version disaggregates the items of the original 12-item instrument, in which different life events were assessed in single items. In the current study, we used the occurrence of any life event in the previous 6 months as a predictor. The 12-item LTE has been shown to have high test-retest reliability (Cohen’s κ >0.77 for all but one item), sensitivity (0.89) and specificity (0.74).^20^

The State-Trait Anxiety Inventory (STAI) measures anxiety at the current moment and in general, addressing each with 20 items.^21^ Respondents score on a Likert scale with categories 1 (“not at all”), 2 (“somewhat”), 3 (“moderately so”), and 4 (“very much so”) for state anxiety and 1 (“almost never”), 2 (“sometimes”), 3 (“often”), and 4 (“almost always”) for trait anxiety. Two subscale sum scores are computed, which have been shown to be internally consistent, with Cronbach’s α coefficients >0.9 for both scales.^22^

The 21-item Beck Depression Inventory (BDI-1A) was administered to measure depression severity.^23^ Items are scored on a 0-4 Likert scale and responses are added up to a sum score. We calculated two subscale scores, in line with previous research.^24^ Items 1–13 (sadness, pessimism, past failure, loss of pleasure, guilty feelings, punishment feelings, self-dislike, self-criticalness, suicidal ideation, crying, agitation, loss of interest, indecisiveness) were summed to a BDI *cognitive* subscore and items 14-21 (body-dislike, activity problems, sleep problems, fatigue, decreased appetite, weight loss, worrying about health, loss of interest in sex) to a BDI *somatic* subscale score. The BDI has good psychometric properties.^23^

The occurrence of a lifetime depressive episode and depression treatment history were assessed by two screening questions in the Pregnancy Risk Questionnaire.^25^

### Somatic and obstetric measures

Hypertension at intake was defined as a systolic blood pressure ≥ 140 mm Hg and/or a diastolic blood pressure ≥ 90 mm Hg. Participants using antihypertensive medication were allocated to the hypertension category. Data on obstetric and somatic history and on medication use at intake were extracted from obstetric records by research assistants under supervision of a medical doctor (EJS). Based on a summary of these data, the risk of complications of pregnancy or delivery was determined by two gynaecologists, who subsequently assigned an obstetric risk score: “normal risk”, “increased risk” or “risk not determinable”. There was fair to moderate agreement between the two observers (Cohen’s κ = 0.39, *P* <0.001). A final obstetric risk score was computed with value “not increased” if both observers scored “not increased”, “increased” if one of the observers scored “increased” and “missing” if one of the observers scored “not determinable”.

### Outcome measures

The following outcome measures were used: gestational age at delivery, birthweight, Apgar score and maternal maximum blood pressure.

In a secondary analysis, outcomes were dichotomised at clinical cut-offs. Preterm birth was defined as delivery before 37 completed weeks of gestation. The cut offs for low and very low birthweight were set at 2500 and 1500 g, respectively (WHO definition). The qualification “small for gestational age” (SGA) was used in case of a birthweight below the 10^th^ centile, using Dutch birthweight charts.^26^ A low Apgar score five minutes after birth was defined as a score below 7. Gestational hypertension was defined as a maximum systolic blood pressure ≥ 140 mm Hg and/or a maximum diastolic blood pressure ≥ 90 mm Hg. Participants using antihypertensive medication were allocated to the hypertension category.

### Missing data

Subjects who did not return questionnaires (n = 13) and one case where pregnancy ended at 25 weeks gestational age because of several foetal congenital malformations were removed from the sample. This resulted in a sample of 584 women, of which 444 had no missing data. In total, 9.6% of the data were missing and imputed 20 times using the Amelia II R-package.^27,28^ Because of collinearity issues the variables intake date, delivery date and birthweight percentile were not included in the imputations.

### Statistical analysis

Analyses were performed on imputed datasets and subsequently pooled. To minimize collinearity in multivariable regression models, Spearman’s rank coefficient (ρ) was calculated to assess correlations between the psychosocial variables. A subset of measures with ρ <|0.7| was included in subsequent analyses.

The residual distributions of univariable regression were examined using Q-Q plots, skewness and kurtosis and showed non-normality. The following transformations were applied to achieve normally distributed model residuals for dependent variables: 7^th^ power for gestational age at delivery, 2^nd^ power for birthweight, and natural logarithm for systolic and diastolic blood pressure. For Apgar score, no transformation was found that led to normally distributed model residuals. Therefore only dichotomised Apgar scores were analysed.

The association between individual somatic, psychological and social variables and each obstetric outcome was investigated using univariable linear and logistic regression. Independent variables with *P* <0.1 in univariable models were included in subsequent multivariable regression. To evaluate added value of psychosocial factors on top of somatic risk factors, we compared explained variance (R^2^) in models with and without psychosocial factors. All statistical procedures were run in IBM SPSS Statistics v25.

## Results

### Descriptive statistics

Table 1 shows the baseline participant characteristics before and after imputation. Main outcomes are displayed in Table 2.

**Table 1.**
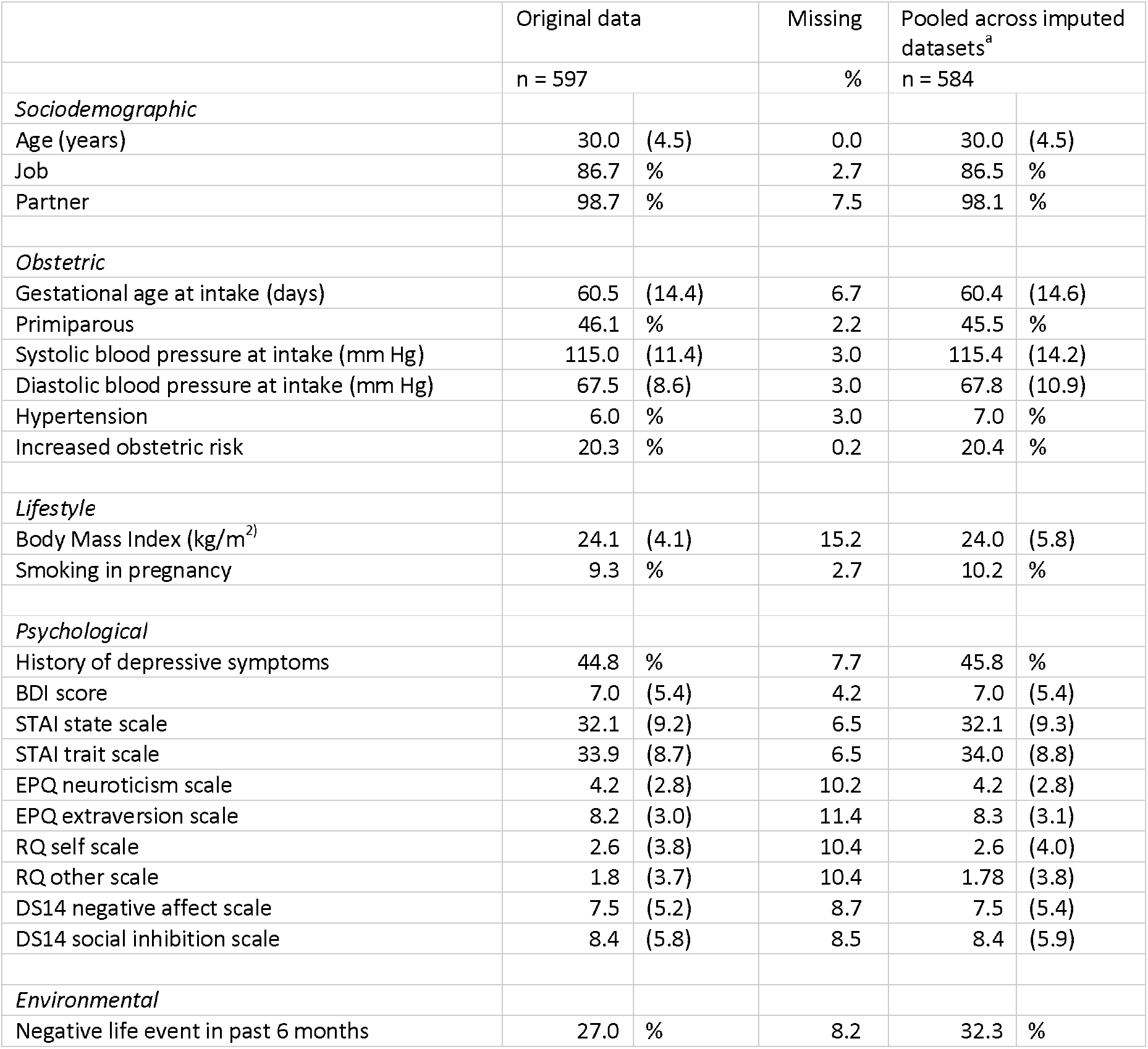
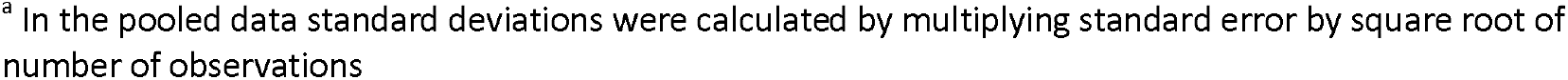
Baseline characteristics of the original study sample and pooled characteristics after multiple imputation Mean (standard deviation) or percentage is displayed

|  | Original data |  | Missing | Pooled across imputed datasets<sup>a</sup> |  |
| --- | --- | --- | --- | --- | --- |
|  | n = 597 |  | % | n = 584 |  |
| *Sociodemographic* |  |  |  |  |  |
| Age (years) | 30.0 | (4.5) | 0.0 | 30.0 | (4.5) |
| Job | 86.7 | % | 2.7 | 86.5 | % |
| Partner | 98.7 | % | 7.5 | 98.1 | % |
| *Obstetric* |  |  |  |  |  |
| Gestational age at intake (days) | 60.5 | (14.4) | 6.7 | 60.4 | (14.6) |
| Primiparous | 46.1 | % | 2.2 | 45.5 | % |
| Systolic blood pressure at intake (mm Hg) | 115.0 | (11.4) | 3.0 | 115.4 | (14.2) |
| Diastolic blood pressure at intake (mm Hg) | 67.5 | (8.6) | 3.0 | 67.8 | (10.9) |
| Hypertension | 6.0 | % | 3.0 | 7.0 | % |
| Increased obstetric risk | 20.3 | % | 0.2 | 20.4 | % |
| *Lifestyle* |  |  |  |  |  |
| Body Mass Index (kg/m<sup>2</sup>) | 24.1 | (4.1) | 15.2 | 24.0 | (5.8) |
| Smoking in pregnancy | 9.3 | % | 2.7 | 10.2 | % |
| *Psychological* |  |  |  |  |  |
| History of depressive symptoms | 44.8 | % | 7.7 | 45.8 | % |
| BDI score | 7.0 | (5.4) | 4.2 | 7.0 | (5.4) |
| STAI state scale | 32.1 | (9.2) | 6.5 | 32.1 | (9.3) |
| STAI trait scale | 33.9 | (8.7) | 6.5 | 34.0 | (8.8) |
| EPQ neuroticism scale | 4.2 | (2.8) | 10.2 | 4.2 | (2.8) |
| EPQ extraversion scale | 8.2 | (3.0) | 11.4 | 8.3 | (3.1) |
| RQ self scale | 2.6 | (3.8) | 10.4 | 2.6 | (4.0) |
| RQ other scale | 1.8 | (3.7) | 10.4 | 1.78 | (3.8) |
| DS14 negative affect scale | 7.5 | (5.2) | 8.7 | 7.5 | (5.4) |
| DS14 social inhibition scale | 8.4 | (5.8) | 8.5 | 8.4 | (5.9) |
| *Environmental* |  |  |  |  |  |
| Negative life event in past 6 months | 27.0 | % | 8.2 | 32.3 | % |

<sup>a</sup> In the pooled data standard deviations were calculated by multiplying standard error by square root of number of observations

**Table 2.** Main outcomes in the study sample, pooled data after multiple imputation Mean (standard deviation) or percentage is displayed n = 569 for gestational age at delivery, n = 568 for small for gestational age, n = 584 for other outcomes

|  |  |  |
| --- | --- | --- |
| *Gestational age at delivery* |  |  |
| Mean (days) | 278 | (13) |
| Preterm delivery (<37 weeks gestation) | 6.9 | % |
| *Birthweight* |  |  |
| Mean (g) | 3560 | (685) |
| Low birthweight (<2500 g) | 5.4 | % |
| Very low birthweight (<1500 g) | 0.6 | % |
| Small for gestational age (<10<sup>th</sup> birthweight centile) | 4.8 | % |
| *Apgar score at 5 minutes* |  |  |
| Mean (points) | 9.6 | (1.2) |
| Low Apgar score (<7) | 2.2 | % |
| *Maternal blood pressure* |  |  |
| Maximum systolic (mm Hg) | 127 | (15) |
| Maximum diastolic (mm Hg) | 79 | (11) |
| Change systolic<sup>a</sup> (mm Hg) | 12 | (15) |
| Change diastolic (mm Hg) | 12 | (10) |
| Hypertension<sup>b</sup> | 28.7 | % |
| De novo hypertension<sup>c</sup> | 23.1 | % |
| Use of antihypertensive medication in pregnancy | 2.1 | % |

<sup>a</sup> Difference between maximum systolic and intake systolic blood pressure
<sup>b</sup> Systolic blood pressure $\geq 140$ mm Hg and/or diastolic blood pressure $\geq 90$ mm Hg and/or use of antihypertensive medication
<sup>c</sup> Normal blood pressure at intake, hypertension later in pregnancy

Correlations were >|0.7| between the BDI and both of its subscales, between STAI trait and STAI state, between STAI trait and DS14 negative affect, and between EPQ and DS14 subscales (S1 Appendix). The following psychosocial measures were included in subsequent analyses: BDI somatic and cognitive, STAI state, RQ self, RQ other, DS14 negative affect, DS14 social inhibition and the LTE. DS14 was chosen over the EPQ because DS14 was related to multiple dependent variables in univariable regression (S2 Appendix).

### Multivariable linear regression (Table 3)

Higher diastolic blood pressure and increased obstetric risk at intake were significantly associated with lower gestational age at delivery. Primiparity and smoking in pregnancy were associated with lower birthweight. High blood pressure at intake was associated with higher maximum diastolic blood pressure. Having a partner, primiparity, high blood pressure, and BMI at intake were associated with higher maximum systolic blood pressure. Looking at psychological variables, only the associations of DS14 negative affect with higher maximum systolic and diastolic blood pressure remained significant in multivariable analyses. Across the 20 imputed datasets, the adjusted R^2^ increased from 0.16-0.28 without to 0.17-0.29 with the DS14 scale when predicting maximum systolic blood pressure (i.e. ΔR^2^ ~ 0.01) and from 0.26-0.29 without to 0.28-0.32 with the DS14 scale when predicting maximum diastolic blood pressure (i.e. ΔR^2^ ~ 0.02-0.03).

**Table 3.**
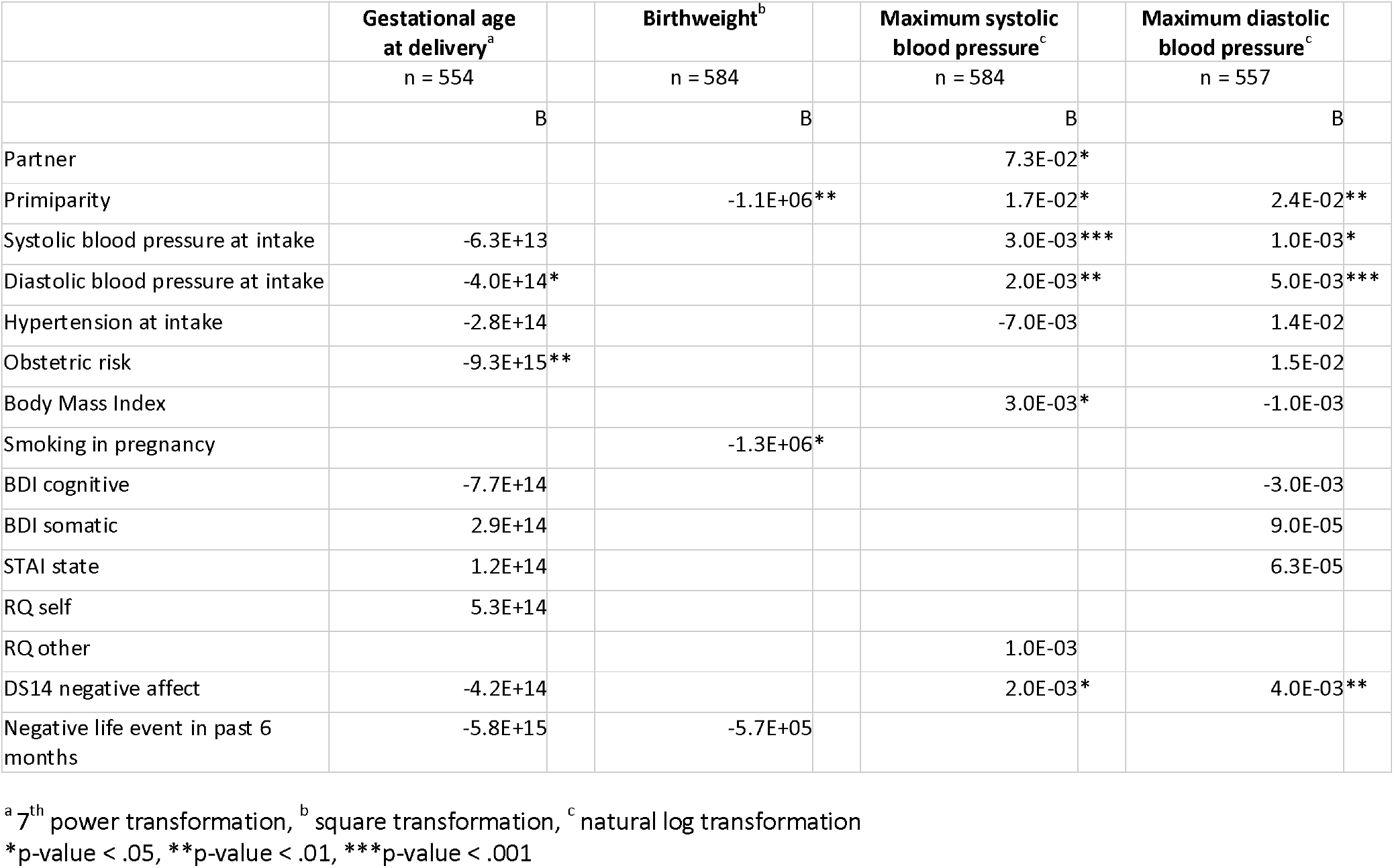
Multivariable linear regression analyses for somatic, psychological and social variables with four obstetric outcomes Pooled data after multiple imputation, variables with *P* <0.1 for univariable association with the outcome of interest

|  | Gestational age<br>at delivery<sup>a</sup> | Birthweight<sup>b</sup> | Maximum systolic<br>blood pressure<sup>c</sup> | Maximum diastolic<br>blood pressure<sup>c</sup> |
| --- | --- | --- | --- | --- |
|  | n = 554 | n = 584 | n = 584 | n = 557 |
|  | B | B | B | B |
| Partner |  |  | 7.3E-02* |  |
| Primiparity |  | -1.1E+06** | 1.7E-02* | 2.4E-02** |
| Systolic blood pressure at intake | -6.3E+13 |  | 3.0E-03*** | 1.0E-03* |
| Diastolic blood pressure at intake | -4.0E+14* |  | 2.0E-03** | 5.0E-03*** |
| Hypertension at intake | -2.8E+14 |  | -7.0E-03 | 1.4E-02 |
| Obstetric risk | -9.3E+15** |  |  | 1.5E-02 |
| Body Mass Index |  |  | 3.0E-03* | -1.0E-03 |
| Smoking in pregnancy |  | -1.3E+06* |  |  |
| BDI cognitive | -7.7E+14 |  |  | -3.0E-03 |
| BDI somatic | 2.9E+14 |  |  | 9.0E-05 |
| STAI state | 1.2E+14 |  |  | 6.3E-05 |
| RQ self | 5.3E+14 |  |  |  |
| RQ other |  |  | 1.0E-03 |  |
| DS14 negative affect | -4.2E+14 |  | 2.0E-03* | 4.0E-03** |
| Negative life event in past 6 months | -5.8E+15 | -5.7E+05 |  |  |

<sup>a</sup> 7<sup>th</sup> power transformation, <sup>b</sup> square transformation, <sup>c</sup> natural log transformation
\*p-value < .05, \*\*p-value < .01, \*\*\*p-value < .001

### Multivariable logistic regression (Table 4)

Primiparity and obstetric risk were each associated with a more than 3-fold increase in the odds of preterm delivery. Higher diastolic blood pressure at intake was associated with slightly higher odds of preterm delivery. Only smoking in pregnancy was associated with higher odds of SGA. Diastolic blood pressure and hypertension at intake and DS14 negative affect were associated with increased odds of gestational hypertension. Across imputed datasets, addition of DS14 negative affect to the model increased adjusted R^2^ estimates from 0.19-0.21 without the DS14 scale to 0.20-0.22 with the DS14 scale (i.e. ΔR^2^ ~ 0.01).

**Table 4.**
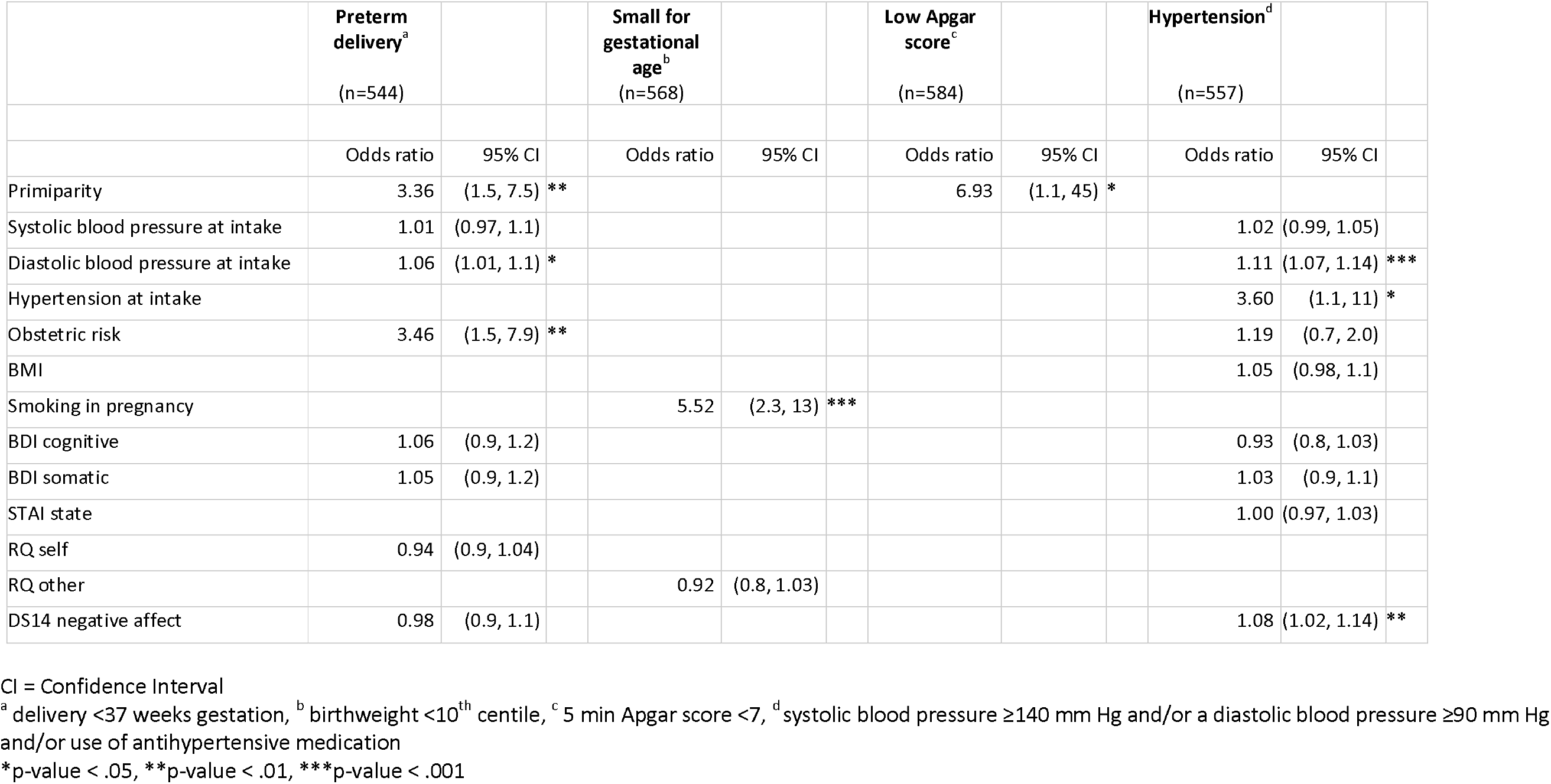
Multivariable logistic regression analyses for somatic, psychological and social variables with four obstetric outcomes Pooled data after multiple imputation, variables with P <0.1 for univariable association with the outcome of interest

## Discussion

The current study investigated the association between pregnancy outcome and a range of obstetric, lifestyle and psychosocial factors measured before 20 weeks gestation. We used routine measurements from obstetric records and added questionnaires on psychosocial items, thus evaluating a low burden screening procedure. In multivariable analyses, obstetric and lifestyle variables measured before 20 weeks of gestation showed independent associations with a range of pregnancy outcomes, whereas psychosocial variables did not, except for negative affect being associated with maternal blood pressure. The current finding of limited predictive value of psychosocial measurements for obstetric outcomes aligns with previous work that included similar covariates.^29,30^ However, other studies did find associations between psychosocial factors and pregnancy outcomes. Some of these studies differ from the current study in that they did not adjust for somatic risk factors.^31,32^ Furthermore, studies that found significant associations between maternal anxiety or depression and pregnancy outcome were performed in developing countries or in mainly low income women,^31,33–37^ some had a retrospective design,^31^ some measured psychosocial factors in the second half of pregnancy,^31,34–36,38,39^ and/or used non-validated measures of risk factors.^33–40^

It is safe to state that findings on the predictive value of psychosocial factors for pregnancy outcome have been mixed so far. A plausible explanation for the varied results could be confounding. It is well known that social adversity, physical disease and mental state are interrelated. For example, depression is more commonly reported in people with diabetes and anxiety is associated with an increased incidence of cardiovascular disease.^41,42^ Thus far, no conclusive evidence has been provided for the exact chronology in pathogenesis of comorbid mental and somatic diseases and it is likely that the association is bidirectional.^43^ Thus, to accurately determine the independent effects of psychosocial risk factors in pregnancy, main obstetric and somatic risk factors should be included in analyses. Without accounting for these risk factors we cannot be sure if and to what extent observed associations between psychosocial factors and pregnancy outcome are confounded, mediated or moderated by somatic factors. Another explanation for inconsistent results could lie in the time between assessments and birth. Increased scores on psychometric measures in late pregnancy might reflect actual symptoms of pregnancy complications instead of anxiety or depression.^44^

The population in the current study consisted mainly of healthy women, mostly employed and with partner. This might explain the low incidence of low birthweight (5.4%), as maternal health and socioeconomic status are known to be related to birthweight.^8^ Incidence of gestational hypertension on the other hand was high (28.7%). This can be explained by the fact that hypertension was assessed based on maximum blood pressure values in obstetric records. These frequently are based on onetime measurements, which makes them sensitive to random fluctuations, unlike in studies where gestational hypertension diagnosis is based on two measurement occasions at least 4 hours apart. Although the study has several strengths, including the broad selection of measurements, prospective design and analytical approach, there were also limitations. First, 43.8% of women asked to participate declined, which may have led to selection bias (e.g., those reluctant to report on mental problems were less likely to be included). Second, the low incidence of birth complications may have limited the statistical power to detect small but relevant predictive associations. Third, data on somatic parameters were derived from questionnaires and obstetric records from different centres with different patient-record systems. Standardised methods could have yielded more reliable data.

## Conclusion

This study supports screening of pregnant women through history taking and physical measurements as already routinely applied in obstetrical practice. Although questionnaires on psychosocial factors are valuable to monitor maternal mental state and overall well-being, the current results indicate that they do not improve identification of women at risk for obstetric complications in a low-risk population of pregnant women.

## Data Availability

The datasets used and analysed during the current study are available from the corresponding author on reasonable request.

## Abbreviations

BDI: Beck Depression Inventory
BMI: Body Mass Index
CI: Confidence Interval
DS14: Type D scale-14
EPQ: Eysenck Personality Questionnaire
LTE: List of Threatening Experiences
OR: Odds Ratio
RQ: Relationship Questionnaire
SGA: Small for Gestational Age
STAI: State-Trait Anxiety Inventory

## Declarations

Ethics approval and consent to participate

All methods were performed in accordance with the relevant guidelines and regulations. The study was approved by the medical research ethics committees (Regionale Toetsingscommissie Patientgebonden Onderzoek MCL and Medisch Ethische Toetsings Commissie VU Medical Centre) of both participating hospitals. All participants gave informed consent.

## Consent for publication

Not applicable

## Authors’ contributions

The statistical analyses were performed and described by KW, RM and EJS. EJS wrote the concept of the remaining text of the manuscript. KW was the first supervisor. He wrote several passages of the final manuscript. AB mainly revised the parts of the manuscript concerning obstetric theory and practice. TO, MW and PJ revised the parts of the text on psychosocial risk factors, their epidemiology and effects on psychic and somatic disease. They also commented on the general design of the study and the manuscript. All authors read and approved the final manuscript.

## Acknowledgements

We would like to thank GGZ Friesland for facilitating the study. We would especially like to thank Erwin Geerts, who deceased August 30, 2010 for his contribution to the study design.

## S1 Appendix

Spearman correlations between administered psychosocial measures

Pooled data after multiple imputation

## S2 Appendix

Univariable linear regression analyses for somatic, psychological and social variables with four obstetric outcomes

Pooled data after multiple imputation

