## Supplementary material for "Psychological and social risk factors and pregnancy outcome: a prospective cohort study": Table 4

Multivariable logistic regression analyses for somatic, psychological and social variables with four obstetric outcomes

Pooled data after multiple imputation, variables with *P* <0.1 for univariable association with the outcome of interest

|  | **Preterm delivery**^a^  (n=544) |  |  | **Small for gestational age**^b^  (n=568) |  |  | **Low Apgar score**^c^  (n=584) |  |  | **Hypertension**^d^  (n=557) |  |  |
| --- | --- | --- | --- | --- | --- | --- | --- | --- | --- | --- | --- | --- |
|  | Odds ratio | 95% CI |  | Odds ratio | 95% CI |  | Odds ratio | 95% CI |  | Odds ratio | 95% CI |  |
| Primiparity | 3.36 | (1.5, 7.5) | ****** |  |  |  | 6.93 | (1.1, 45) | ***** |  |  |  |
| Systolic blood pressure at intake | 1.01 | (0.97, 1.1) |  |  |  |  |  |  |  | 1.02 | (0.99, 1.05) |  |
| Diastolic blood pressure at intake | 1.06 | (1.01, 1.1) | ***** |  |  |  |  |  |  | 1.11 | (1.07, 1.14) | ******* |
| Hypertension at intake |  |  |  |  |  |  |  |  |  | 3.60 | (1.1, 11) | ***** |
| Obstetric risk | 3.46 | (1.5, 7.9) | ****** |  |  |  |  |  |  | 1.19 | (0.7, 2.0) |  |
| BMI |  |  |  |  |  |  |  |  |  | 1.05 | (0.98, 1.1) |  |
| Smoking in pregnancy |  |  |  | 5.52 | (2.3, 13) | ******* |  |  |  |  |  |  |
| BDI cognitive | 1.06 | (0.9, 1.2) |  |  |  |  |  |  |  | 0.93 | (0.8, 1.03) |  |
| BDI somatic | 1.05 | (0.9, 1.2) |  |  |  |  |  |  |  | 1.03 | (0.9, 1.1) |  |
| STAI state |  |  |  |  |  |  |  |  |  | 1.00 | (0.97, 1.03) |  |
| RQ self | 0.94 | (0.9, 1.04) |  |  |  |  |  |  |  |  |  |  |
| RQ other |  |  |  | 0.92 | (0.8, 1.03) |  |  |  |  |  |  |  |
| DS14 negative affect | 0.98 | (0.9, 1.1) |  |  |  |  |  |  |  | 1.08 | (1.02, 1.14) | ****** |

CI = Confidence Interval

^a^ delivery <37 weeks gestation, ^b^ birthweight <10^th^ centile, ^c^ 5 min Apgar score <7, ^d^ systolic blood pressure ≥140 mm Hg and/or a diastolic blood pressure ≥90 mm Hg and/or use of antihypertensive medication

*p-value < .05, **p-value < .01, ***p-value < .001
