## Appendix 1 for "Psychological and social risk factors and pregnancy outcome: a prospective cohort study"

**S1 Appendix**

Spearman correlations between administered psychosocial measures, pooled data after multiple imputation

n = 584

Coefficients >|0.7| are printed in bold font

| Independent variable | BDI | BDI cog | BDI som | STAI state | STAI trait | EPQ-NE | EPQ-EX | RQSM | RQOM | DS14-NA | DS14-SI | Life event |
| --- | --- | --- | --- | --- | --- | --- | --- | --- | --- | --- | --- | --- |
| BDI | 1 |  |  |  |  |  |  |  |  |  |  |  |
| BDI cognitive | **0.88** | 1 |  |  |  |  |  |  |  |  |  |  |
| BDI somatic | **0.90** | 0.61 | 1 |  |  |  |  |  |  |  |  |  |
| STAI state | 0.61 | 0.58 | 0.51 | 1 |  |  |  |  |  |  |  |  |
| STAI trait | 0.61 | 0.63 | 0.48 | **0.71** | 1 |  |  |  |  |  |  |  |
| EPQ neuroticism | 0.50 | 0.53 | 0.39 | 0.48 | 0.68 | 1 |  |  |  |  |  |  |
| EPQ extraversion | -0.12 | -0.13 | -0.10 | -0.14 | -0.21 | -0.25 | 1 |  |  |  |  |  |
| RQ self model | -0.29 | -0.31 | -0.21 | -0.34 | -0.40 | -0.40 | 0.26 | 1 |  |  |  |  |
| RQ other model | -0.07 | -0.08 | -0.06 | -0.15 | -0.19 | -0.18 | 0.28 | 0.14 | 1 |  |  |  |
| DS14 negative affect | 0.49 | 0.51 | 0.37 | 0.54 | **0.72** | **0.77** | -0.21 | -0.36 | -0.13 | 1 |  |  |
| DS14 social inhibition | 0.21 | 0.22 | 0.18 | 0.26 | 0.32 | 0.34 | **-0.71** | -0.32 | -0.32 | 0.36 | 1 |  |
| Negative life event | 0.11 | 0.13 | 0.07 | 0.11 | 0.09 | 0.07 | 0.12 | -0.02 | -0.02 | 0.08 | -0.04 | 1 |
