## Appendix 2 for "Psychological and social risk factors and pregnancy outcome: a prospective cohort study"

**S2 Appendix**

Univariable linear regression analyses for somatic, psychological and social variables with four obstetric outcomes

Pooled data after multiple imputation

*P* values <0.1 are printed in bold font

|  | **Gestational age at delivery** |  | **Birthweight** |  | **Apgar score**  **at 5 min** |  | **Maximum systolic**  **blood pressure** |  | **Maximum diastolic blood pressure** |  |
| --- | --- | --- | --- | --- | --- | --- | --- | --- | --- | --- |
| Transformation | 7th power |  | square |  | none |  | natural log |  | natural log |  |
| Regression | Linear |  | Linear |  | Logistic |  | Linear |  | Linear |  |
|  | B | *P* | B | *P* | OR | *P* | B | *P* | B | *P* |
| Age | -2.0E+14 | 0.52 | 1.8E+4 | 0.65 | 1.00 | 0.94 | 0.001 | 0.17 | 0.001 | 0.62 |
| Job | 6.6E+15 | 0.12 | -1.1E+5 | 0.83 | 0.81 | 0.79 | 0.001 | 0.95 | -0.001 | 0.93 |
| Partner | 1.9E+15 | 0.87 | 6.2E+5 | 0.68 | 22E+5 | 1.00 | 0.074 | **0.03** | -0.005 | 0.89 |
| Primiparity | -3.9E+13 | 0.99 | -1.1E+6 | **0.00** | 6.93 | **0.04** | 0.017 | **0.08** | 0.019 | **0.09** |
| Systolic BP at intake | -2.5E+14 | **0.04** | -1.3E+4 | 0.55 | 1.01 | 0.87 | 0.004 | **0.00** | 0.004 | **0.00** |
| Diastolic BP at intake | -5.3E+14 | **0.00** | -2.2E+4 | 0.42 | 1.00 | 0.96 | 0.004 | **0.00** | 0.007 | **0.00** |
| Hypertension at intake | -1.1E+16 | **0.07** | 1.0E+6 | 0.56 | 0.00 | 1.00 | 0.103 | **0.00** | 0.134 | **0.00** |
| Obstetric risk | -1.2E+16 | **0.00** | -6.7E+5 | 0.12 | 0.42 | 0.40 | 0.017 | 0.12 | 0.034 | **0.01** |
| BMI | -1.4E+14 | 0.72 | 6.8E+4 | 0.19 | 1.04 | 0.55 | 0.006 | **0.00** | 0.006 | **0.00** |
| Smoking in pregnancy | -7.9E+15 | 0.10 | -1.4E+6 | **0.02** | 1.75 | 0.47 | -0.024 | 0.15 | -0.018 | 0.32 |
| Previous smoking^a^ | 5.3E+15 | 0.21 | -2.0E+5 | 0.70 | 1.31 | 0.74 | -0.003 | 0.78 | 0.005 | 0.69 |
| History of depressive symptoms | -3.9E+15 | 0.19 | -4.0E+5 | 0.28 | 0.84 | 0.77 | 0.006 | 0.53 | 0.011 | 0.27 |
| Treatment for depressive symptoms^b^ | 3.0E+13 | 1.00 | -2.9E+5 | 0.60 | 0.25 | 0.23 | 0.016 | 0.28 | 0.015 | 0.39 |
| BDI | -7.6E+14 | **0.00** | -1.2E+4 | 0.73 | 1.01 | 0.84 | 0.001 | 0.12 | 0.002 | **0.04** |
| BDI cognitive | -1.4E+15 | **0.00** | -2.6E+4 | 0.65 | 1.03 | 0.67 | 0.002 | 0.20 | 0.003 | **0.09** |
| BDI somatic | -9.3E+14 | **0.07** | -9.7E+3 | 0.88 | 0.99 | 0.91 | 0.003 | 0.12 | 0.004 | **0.05** |
| STAI state | -2.8E+14 | **0.07** | -2.2E+4 | 0.25 | 0.99 | 0.77 | 0.001 | 0.27 | 0.001 | **0.01** |
| STAI trait | -4.7E+14 | **0.00** | -2.7E+4 | 0.20 | 0.98 | 0.65 | 0.001 | **0.07** | 0.002 | **0.00** |
| EPQ neuroticism | -1.2E+15 | **0.02** | -4.3E+4 | 0.51 | 0.98 | 0.85 | 0.002 | 0.26 | 0.003 | 0.10 |
| EPQ extraversion | -4.1E+14 | 0.40 | -1.4E+4 | 0.82 | 1.00 | 0.98 | 0.002 | 0.14 | 0.002 | 0.29 |
| RQ self | 8.2E+14 | **0.03** | 2.6E+4 | 0.59 | 1.13 | 0.18 | 0.000 | 0.86 | -0.001 | 0.58 |
| RQ other | 6.2E+14 | 0.11 | 6.9E+4 | 0.16 | 0.97 | 0.71 | 0.002 | **0.08** | 0.001 | 0.57 |
| DS14 negative affect | -7.3E+14 | **0.01** | -3.9E+4 | 0.26 | 0.93 | 0.27 | 0.002 | **0.05** | 0.003 | **0.00** |
| DS14 social inhibition | 1.2E+14 | 0.63 | 2.3E+3 | 0.94 | 0.99 | 0.83 | -0.001 | 0.21 | -0.001 | 0.23 |
| Negative life event in past 6 months | -7.1E+15 | **0.04** | -7.8E+5 | **0.08** | 0.64 | 0.58 | 0.005 | 0.58 | 0.010 | 0.41 |

^a^ previous smoking vs. non-smoking

^b^ depressive symptoms with treatment vs. without treatment
